# Benchmarking transformer-based models for medical record deidentification: A single centre, multi-specialty evaluation

**DOI:** 10.1101/2025.05.05.25326979

**Authors:** Rachel Kuo, Andrew A.S. Soltan, Ciaran O’Hanlon, Alan Hasanic, David A. Clifton, Collins Gary, Dominic Furniss, David W. Eyre

## Abstract

**Background:** Robust de-identification is necessary to preserve patient confidentiality and maintain public acceptance of electronic health record (EHR) research. Manual redaction of personally identifiable information (PII) outside of structured data is time-consuming and expensive, limiting the scale of data-sharing possible. Automated de-identification (DeID) could alleviate this burden, with competing approaches including task-specific models and generalist large language models (LLMs). We aimed to identify the optimal strategy for PII redaction, evaluating a number of task specific transformer-architecture models and generalist LLMs using no- and low-adaptation techniques.

**Methods:** We evaluated the performance of four task-specific models (Microsoft Azure DeID service, AnonCAT, OBI RoBERTa & BERT i2b2 DeID) and five general-purpose LLMs (Gemma-7b-IT, Llama-3-8B-Instruct, Phi-3-mini-128k-instruct, GPT-3.5-turbo-0125, GPT-4-0125) at de-identifying 3650 medical records from a UK hospital group, split into general and specialised datasets. Records were dual-annotated by clinicians for PII. The primary outcomes were F1 score, precision, and recall for each comparator in classifying words as PII vs. non-PII. The secondary outcomes were performance per-PII-subtype per-dataset, and the Levenshtein distance as a proxy for hallucinations/addition of extra text. We report untuned performance for task-specific models and zero-shot performance for LLMs. To assess sensitivity to data shifts between hospital sites, we undertook concept alignment and fine-tuning of one task-specific model (AnonCAT), and performed few-shot (1, 5, and 10) in-context learning for each LLM using site-specific data.

**Results:** 17496/479760 (3.65%) words were PII. Inter-annotator F1 for word-level PII was 0.977 (95%CI 0.957-0.991). The best performing redaction tool was the Microsoft Azure de-identification service: F1 0.939 (0.934-0.944), precision 0.928 (0.922-0.934), recall 0.950 (0.943-0.958). The next-best tools were fine-tuned-AnonCAT: F1 0.910 (0.905-0.914), precision 0.978 (0.973-0.982), recall 0.850 (0.843-0.858), and GPT-4-0125 (ten-shots): F1 0.898 (0.876-0.915), precision 0.874 (0.834-0.906), recall 0.924 (0.914-0.933). There was hallucinatory output in Phi-3-mini-128k-instruct and Llama-3-8B-Instruct at zero-, one-, and five-shots, and universally for Gemma-7b-IT. AnonCAT showed significant improvement in performance on fine-tuning (F1 increase from 0.851; 0.843-0.859 to 0.910; 0.905-0.914). Names/dates were consistently redacted by all comparators; there was variable performance for other categories. Fine-tuned-AnonCAT demonstrated the least performance shift across datasets.

**Conclusion:** Automated EHR de-identification using transformer models could facilitate large-scale, domain-agnostic record sharing for medical research alongside other safeguards to prevent reidentification. Low-adaptation strategies may improve the performance of generalist LLMs and task-specific models.

## Introduction

Routinely collected healthcare data for millions of patients are stored within electronic health records (EHR) to facilitate care delivery and efficient communication between professionals^1,2^. These data are diverse and include structured data, such as laboratory results, and semi-structured or unstructured information, such as diagnostic reports. The resulting data-store is a valuable resource for research, audit, education and quality improvement^3^. Recently, there has been increasing interest in utilising EHR data to develop and validate deep learning models to improve clinical outcomes and care eficiency^4^.

Sharing data for secondary use necessitates robust de-identification to preserve confidentiality. Identifiers, such as patient names, must be removed to minimize the risk of re-identification, comply with data protection legislation, and establish public trust and acceptability^5,6^. In Europe, General Data Protection Regulation defines personal data as any information that relates to an identified or identifiable individual. In the US, identified healthcare records are personally identifiable information (PII) when they contain any of 18 identifiers defined in the Health Insurance Portability and Accountability Act (HIPAA).

Effective redaction of PII from semi-or un-structured medical records can be challenging. Manual de-identification, in which humans review and redact personal information, is time-consuming and costly, limiting the scale of data that can safely be made available^7^. Automated de-identification offers considerable benefits to hospitals, researchers, and patients. However, some categories of PII, such as names and locations are poorly recognised using regular expressions or rule-based algorithms, requiring curation of extensive local dictionaries to ensure adequate redaction^8–10^. Rule-based models may be prone to over-redaction or fail to account for nuanced differences in regional formats, compromising the utility of de-identified data^8,9^. Strategies relying on local dictionaries are vulnerable to spelling errors and require re-definition if applied to new datasets^10^. Additionally, there is heterogeneity in clinical language across medical specialties and sites, necessitating domain adaptation to maintain performance.

Domain-agnostic automated de-identification that requires no or minimal adaptation could facilitate safe and cost-effective data sharing at scale. Recently, transformer-based natural language processing (NLP) models have shown promise in de-identification without the requirement for domain-specific adaptation^9,11,12^. Existing research shows that LLMs may be able to perform clinical/biomedical NLP tasks with no domain-specific training (zero-shot inference), or with a few examples of input and desired output (few-shot learning)^9,13,14^. A critical challenge of LLMs is the phenomenon of ’hallucination’, in which LLMs generate erroneous output; e.g., offering opinions on the reference text^15–17^. However, it is unclear how large generalist models perform in comparison to task-specific models. Moreover, some task-specific models permit adaptation and fine-tuning, but the quantitative benefit of this for deidentification is not yet clear.

In this study, we evaluated four existing proprietary de-identification software tools, and five LLMs in the task of text de-identification, using no- and minimal-adaptation strategies, and a new dataset of 3650 mixed clinical records from a group of National Health Service (NHS) hospitals in the United Kingdom.

## Methods

### Dataset

We used routinely collected data from Oxford University Hospitals NHS Foundation Trust (OUHNFT), a large teaching trust comprised of four hospitals providing care to around 1% of the UK population as well as specialist referral services^18^. Data were obtained from two sources between January-2020-January-2022 inclusive. We randomly sampled 1000 general radiology and 1000 histopathology reports from the Infections in Oxfordshire Research Database (IORD). IORD has approvals from the National Research Ethics Service South Central – Oxford C Research Ethics Committee (19/SC/0403), the Health Research Authority and the Confidentiality Advisory Group (19/CAG/0144). Structured identifiers were removed from records prior to analysis, but free-text was provided without further redaction to researchers with NHS contracts for analysis within NHS infrastructure. We also randomly sampled 550 radiograph, 550 computed tomography (CT), and 550 magnetic resonance (MR) request and report entries from a specialised musculoskeletal database, with Camden and Kings Cross Research Ethics Committee approval (22/LO/0049).

### PII definition and annotation rules

We developed our labelling ontology based on HIPAA PII categories^19^, including three further categories we considered additional potential identifiers. These were: hospital/unit names, private healthcare organisation names, and individual professional details where included as part of a healthcare professionals’ name or where related to patients (Table 1). We excluded two inapplicable HIPAA categories (biometric identifiers and full-face photographic images).

**Table 1.** Categories of PII.

|  | **PII category** | **PII subcategory** |
| --- | --- | --- |
| **1** | Names | Patient or relative |
|  |  | Healthcare worker |
| **2** | Address | House or street number |
|  |  | Street name |
|  |  | City |
|  |  | County |
|  |  | Country |
|  |  | Postcode |
| **3** | Dates | Dates |
|  |  | Age over 89 years |
| **4** | Phone numbers |  |
| **5** | Fax numbers |  |
| **6** | Electronic email addresses |  |
| **7** | Social security numbers |  |
| **8** | Medical record numbers | Medical record number |
|  |  | NHS number |
|  |  | Specimen identifier |
| **9** | Health plan beneficiary numbers |  |
| **10** | Account numbers |  |
| **11** | Certificate/license numbers | GMC number |
|  |  | NMC number |
|  |  | Any other license |
| **12** | Vehicle identifiers and serial numbers |  |
| **13** | Device identifiers and serial numbers |  |
| **14** | Web Universal Resource Locators (URLs) |  |
| **15** | Internet Protocol (IP) address numbers |  |
| **16** | Any other unique identifying number,<br>characteristic, or code | Hospital/unit |
|  |  | External healthcare organisation |
|  |  | Professional details |

All examples were annotated by two clinically qualified labellers (two of RK, CO, and AH), using the open-source software *doccano*, hosted on an OUHNFT server^20^. Annotations were applied on a word-level and included PII category, and the start and end position of each annotation. After completion of labelling, any disagreements were recorded, and then resolved with discussion.

### Deidentification models

We evaluated models using no, or minimal, adaptation approaches for de-identification including four purpose-built de-identification software based upon the transformer architecture (Microsoft Azure de-identification service, AnonCAT, One Brave Idea [OBI] RoBERTa & BERT i2b2 Models^21,22^) and five LLMs (Gemma-7b-IT, Llama-3-8B-Instruct, Phi-3-mini-128k-instruct, GPT-3.5-turbo-0125 and GPT4-0125^21–25^; details in Supplement).

For each LLM, we compared performance with zero, one-, five- and ten-shot learning, primed using examples sourced from outside the dataset (Supplement). Task-specific models were evaluated as provided by the manufacturers (Supplement). All 3650 examples were used for evaluating performance.

To assess the change in performance brought about by adaptation of one tuneable task-specific model, we evaluated AnonCAT both using its default weights and updated weights following concept-expansion and fine-tuning. A randomly selected sample of 365 (10%) clinical notes stratified by dataset was used for tuning with a learning rate of 0.00002 over 10 epochs (Supplement), and the tuned-model was evaluated used the remaining 3285 documents.

We considered that some, but not all, uses of secondary data would necessitate the redaction of professional titles where this was related to healthcare professionals. The default behaviours of the Microsoft Azure de-identification service and OBI models was to redact professional titles. However, by default, the AnonCAT model does not redact professional titles and we therefore performed additional analyses of the untuned model to report performance without the requirement to redact professional titles. During adaptation, we expanded the concept set of the model to align AnonCAT’s base ontology with our own (adding concepts for professional titles, external healthcare organisation, hospital/unit, age over 89, account, URL, GMC number, and social security number). We therefore applied the requirement to redact professional titles only to the tuned model, i.e., similar to all other models in this study.

All inference, training, and analyses were performed on a virtual machine hosted on OUHNFT’s Azure tenancy between April and June 2024.

### Prompt structure

Optimising LLM prompts improves downstream task performance^26^. We provided each LLM with a structured prompt. First, we specify the task (‘Anonymise the following text’). Second, we specify rules for task completion (’Replace all identifiers with their classification’). Third, we provide the labelling ontology, and a brief, synthetic example of desired output (‘If you find a doctor’s name, such as ‘Dr John Doe’, replace this with [profession] [doctor] [doctor]’). Finally, we describe undesirable output (’Do not output any additional text’).

### Statistical analysis and evaluation metrics

We estimate word-level inter-annotator reliability of clinicians using precision (positive predictive value of a PII label), recall (sensitivity, the proportion of all PII detected) and pairwise F1 scores (harmonic mean of precision and recall), using two prediction classes (PII/non-PII)^27^. To assess variation between clinician-performed de-identification, the most senior clinician (RK) was chosen as the gold standard, and precision/recall were calculated using this assumption. The calculation of pairwise F1 score is independent of the designation of annotators as the gold standard or experimental source^27^.

Model performances were compared to a composite reference standard combining two clinician annotations; reporting precision, recall and F1 score for two prediction classes (PII/non-PII).

To describe per-category recall, we used the clinician-assigned PII categories as a reference and considered the PII to be correctly identified if it was redacted under any model-generated label. This reflects that some PII may be correctly classified in two categories, that proprietary models had different labelling ontologies, and redaction rather than PII classification is most important in real-world settings. We calculate recall for all categories comprising >1% of all PII in the dataset.

We defined LLM hallucinations as additional text that was not present in the reference text. As a proxy, we calculated BLEU scores and Levenshtein distance, commonly used metrics for measuring string similarity^28,29^. The Levenshtein distance is defined as the minimum number of character-edits required to transform one string to another; here, from redacted to reference records^28^. BLEU scores are defined by *n*-gram overlap between two strings; here, we calculated the cumulative 4-gram BLEU score^29^. Low BLEU scores and high Levenshtein distances are potentially indicative of LLM hallucination. We qualitatively examined 50 randomly selected samples of each model output. We calculated 95% confidence intervals (CI) using bootstrapping with 10,000 samples. We coded all analyses using Python (v3.8.10).

## Results

### Dataset description

We analysed 3650 records consisting of 479760 words, of which 17496 (3.65%) were PII. Record length and PII prevalence differed across datasets (Table S1). The most frequent forms of PII were names (6901/17496, 39.4%), of which the majority were healthcare professional names (6870/6901, 99.6%) (Table S2). In common with previous research, only a minority of names were patient names (31, 0.4%)^30^.

The next most frequent PII categories were ‘other unique identifiers’ (4758, 27.2%; comprising of professional details, names of external healthcare organisations, and names of hospitals or healthcare units), dates (3641, 20.8%), medical record numbers (1408, 8.0%), and telephone numbers (334, 1.9%). All other PII categories had a prevalence of <1%. There were no occurrences of fax numbers, health plan beneficiary numbers, vehicle/device identifiers, or IP addresses.

### Inter-annotator results

There was excellent agreement between clinician annotators. Pairwise-F1 for classification of PII/non-PII was 0.977 (0.957-0.991), precision 0.967 (0.932-0.993), and recall 0.986 (0.971-0.997) (Table S3). Character-level Cohen’s Kappa was 0.895 (95% CI: 0.886–0.904), signifying high consistency in how PII was delineated by clinician annotators. All discrepancies between annotators were due to cases in which one annotator did not notice a PII-word. Once identified, there were no disagreements about whether a word should be classified as PII. The BLEU score between the original, unredacted records, and manually redacted records was 0.931 (0.923-0.934), reflecting the prevalence of PII in the dataset (Table 2). The Levenshtein distance was 67.0 (63.3-70.8).

**Table 2.**
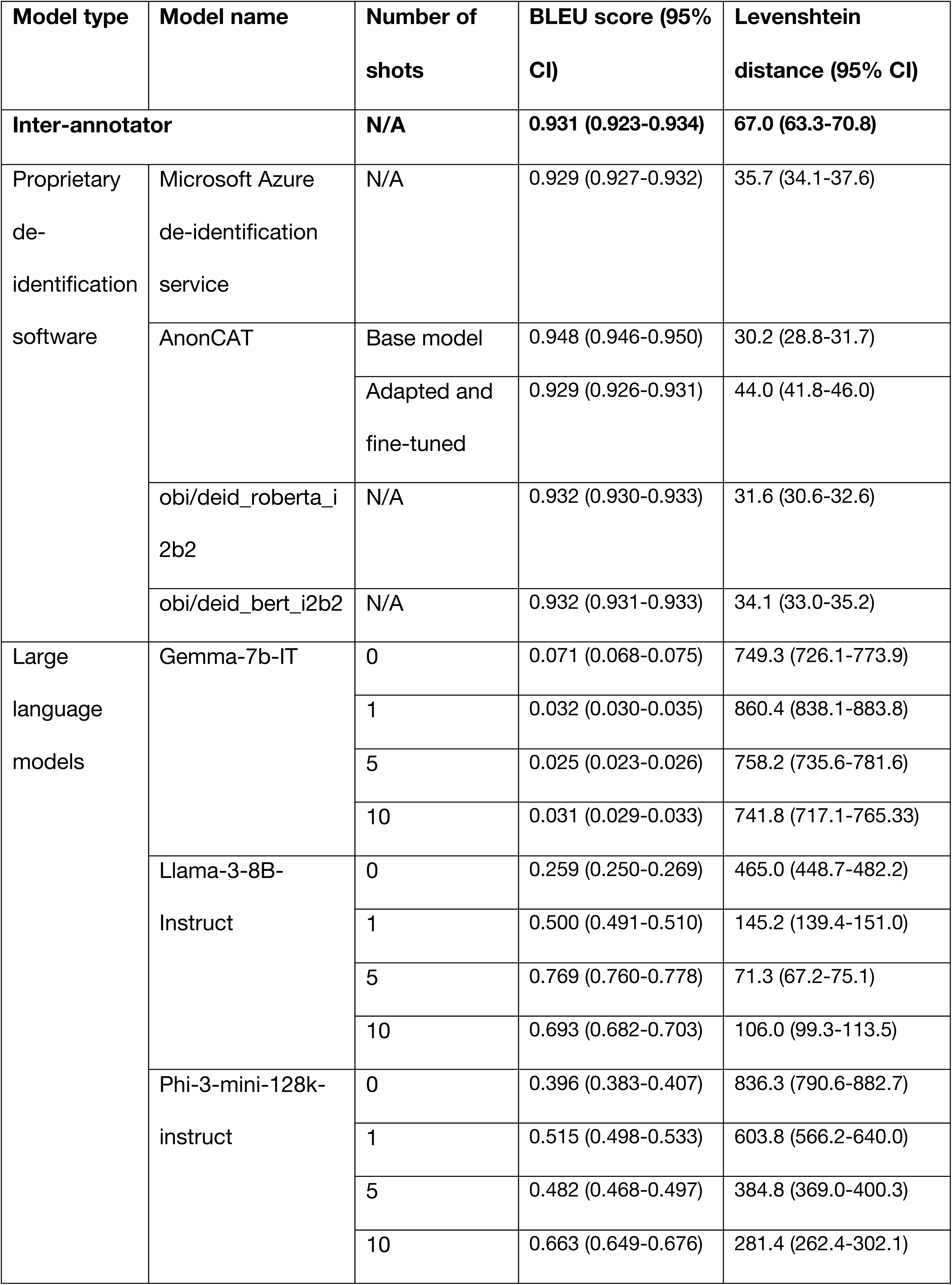
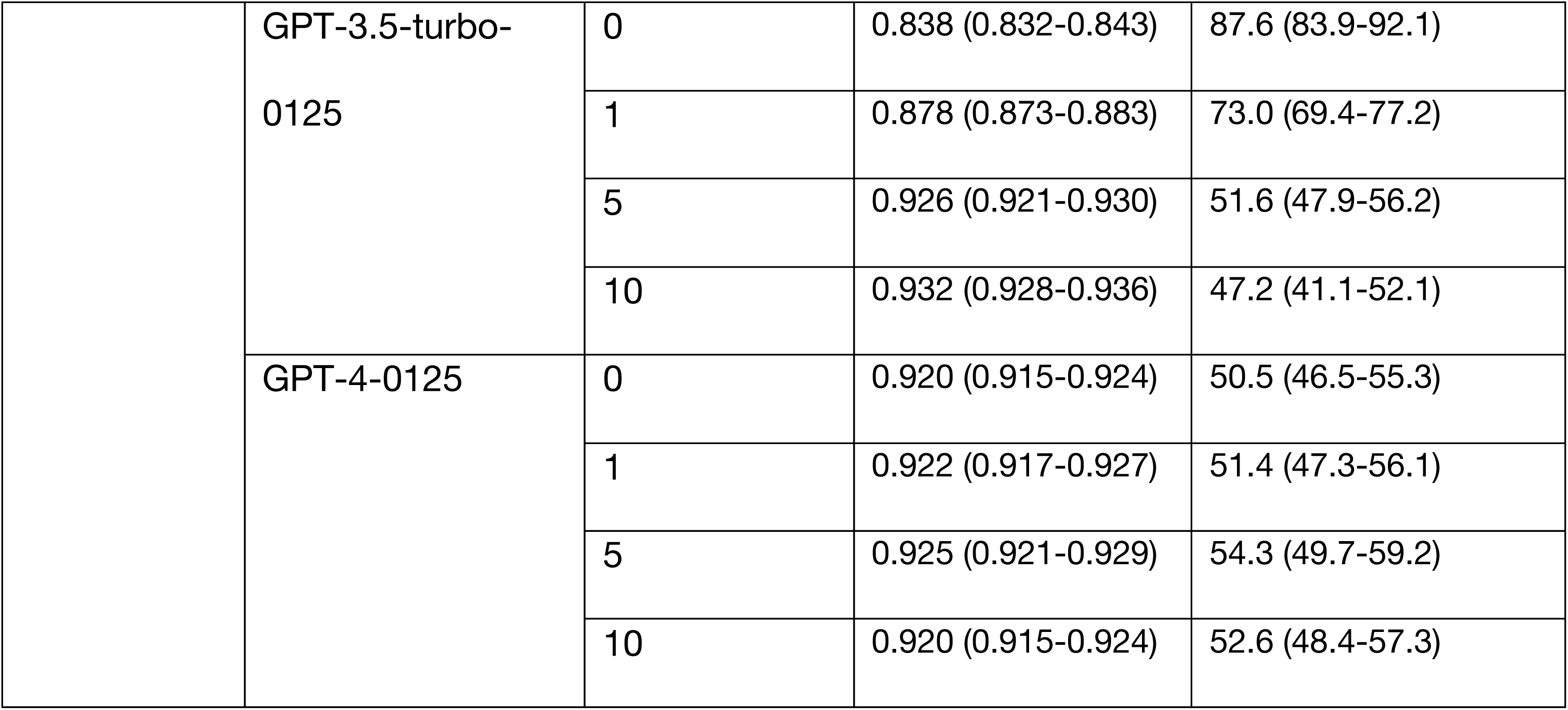
String similarity between original and redacted text. Per model BLEU scores and Levenshtein distances are shown.

| Model type | Model name | Number of shots | BLEU score (95% CI) | Levenshtein distance (95% CI) |
| --- | --- | --- | --- | --- |
| **Inter-annotator** |  | **N/A** | **0.931 (0.923-0.934)** | **67.0 (63.3-70.8)** |
| Proprietary de-identification software | Microsoft Azure de-identification service | N/A | 0.929 (0.927-0.932) | 35.7 (34.1-37.6) |
|  | AnonCAT | Base model | 0.948 (0.946-0.950) | 30.2 (28.8-31.7) |
|  |  | Adapted and fine-tuned | 0.929 (0.926-0.931) | 44.0 (41.8-46.0) |
|  | obi/deid_roberta_i2b2 | N/A | 0.932 (0.930-0.933) | 31.6 (30.6-32.6) |
|  | obi/deid_bert_i2b2 | N/A | 0.932 (0.931-0.933) | 34.1 (33.0-35.2) |
| Large language models | Gemma-7b-IT | 0 | 0.071 (0.068-0.075) | 749.3 (726.1-773.9) |
|  |  | 1 | 0.032 (0.030-0.035) | 860.4 (838.1-883.8) |
|  |  | 5 | 0.025 (0.023-0.026) | 758.2 (735.6-781.6) |
|  |  | 10 | 0.031 (0.029-0.033) | 741.8 (717.1-765.33) |
|  | Llama-3-8B-Instruct | 0 | 0.259 (0.250-0.269) | 465.0 (448.7-482.2) |
|  |  | 1 | 0.500 (0.491-0.510) | 145.2 (139.4-151.0) |
|  |  | 5 | 0.769 (0.760-0.778) | 71.3 (67.2-75.1) |
|  |  | 10 | 0.693 (0.682-0.703) | 106.0 (99.3-113.5) |
|  | Phi-3-mini-128k-instruct | 0 | 0.396 (0.383-0.407) | 836.3 (790.6-882.7) |
|  |  | 1 | 0.515 (0.498-0.533) | 603.8 (566.2-640.0) |
|  |  | 5 | 0.482 (0.468-0.497) | 384.8 (369.0-400.3) |
|  |  | 10 | 0.663 (0.649-0.676) | 281.4 (262.4-302.1) |
|  | GPT-3.5-turbo-0125 | 0 | 0.838 (0.832-0.843) | 87.6 (83.9-92.1) |
|  |  | 1 | 0.878 (0.873-0.883) | 73.0 (69.4-77.2) |
|  |  | 5 | 0.926 (0.921-0.930) | 51.6 (47.9-56.2) |
|  |  | 10 | 0.932 (0.928-0.936) | 47.2 (41.1-52.1) |
|  | GPT-4-0125 | 0 | 0.920 (0.915-0.924) | 50.5 (46.5-55.3) |
|  |  | 1 | 0.922 (0.917-0.927) | 51.4 (47.3-56.1) |
|  |  | 5 | 0.925 (0.921-0.929) | 54.3 (49.7-59.2) |
|  |  | 10 | 0.920 (0.915-0.924) | 52.6 (48.4-57.3) |

## Model results

### PII vs. non-PII

There was substantial variation in performance between the evaluated models (Figure 1, Table S3). The Microsoft Azure de-identification service had the highest F1 score 0.939 (95% CI 0.934-0.944), precision of 0.928 (0.922-0.934), and recall of 0.950 (0.943-0.958), approaching clinician performance. The fine-tuned, concept-expanded AnonCAT (FT-AnonCAT) model had an F1 score 0.910 (0.905-0.914), precision 0.978 (0.973-0.982) and recall 0.850 (0.843-0.858), including the redaction of healthcare professional titles as this was added during the adaptation process.

**Figure 1.**
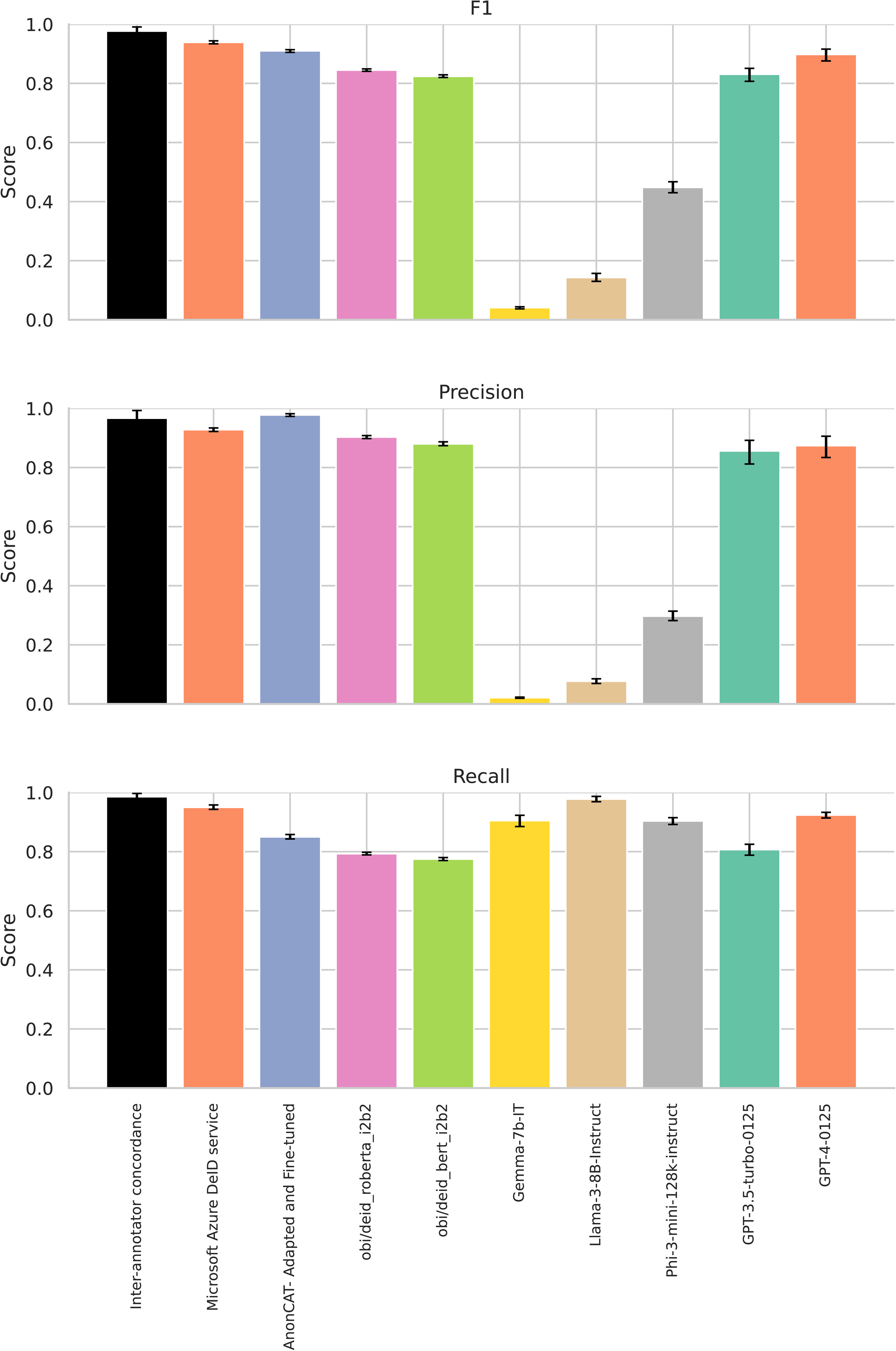
Model performance, for PII vs. non-PII classification, with 95% confidence intervals. All LLM results are plotted using performance with ten-shots. FT-AnonCAT performance is plotted with the requirement to redact healthcare professional titles. **Panel A shows F1 score, panel B recall, and panel C precision.**

The OBI RoBERTa i2b2 model outperformed the OBI BERT i2b2 model, possibly reflecting strengths of the underlying RoBERTa model’s training procedure, including a larger training corpus and batch size, dynamic masking procedure, and improved hyperparameter optimisation. OBI RoBERTA i2b2 outperformed GPT-3.5-turbo-0125 with 10 shot prompting as measured by F1 score, but was surpassed by GPT-4-0125.

The best performing LLM was GPT-4-0125 with ten-shot learning, with F1 score 0.898 (0.876-0.915), precision 0.874 (0.834-0.906), and recall 0.924 (0.914-0.933; Figure S1). This was followed by GPT-3.5-turbo-0125 with ten-shot learning, with F1 score 0.831 (0.807-0.851), precision 0.856 (0.812-0.892), and recall 0.807 (0.788-0.825).

There was improvement in GPT-3.5-turbo-0125 with few-shot learning: the F1 score rose from 0.530 (0.514-0.547) at zero shots to 0.831 (0.807-0.851) with 10 shots, driven by improved precision; recall remained similar through all iterations of zero- and few-shot learning. On qualitative examination with none, or fewer in-context examples, the LLM over-redacted records, including clinically relevant information such as diagnosis, or details of pathology.

The performance of other LLMs was more modest. The next best was Phi-3-mini-128k-instruct, also improving with few-shot learning, with F1 score 0.146 (0.140-0.153) at zero-shots, improving to 0.448 (0.430-0.467) with ten-shots. However, there was significant imbalance between precision and recall. At ten-shots, precision was 0.297 (0.282-0.314) and recall 0.904 (0.892-0.915). This was consistent with our findings on qualitative examination, showing over-redacted records.

Llama-3-8B-Instruct showed the same pattern of over-redaction, showing best performance at five-shots, with an F1 0.198 (0.181-0.216), precision 0.077 (0.069-0.085) and recall 0.990 (0.983-0.995). We did not observe any change in performance from zero-to few-shot learning with Gemma-7b-IT. The F1 at zero-shots was 0.089 (0.086-0.092), and at ten-shots 0.041 (0.037-0.044). At 10-shots, precision was 0.021 (0.019-0.023) and recall was 0.905 (0.885-0.923). Qualitative examination Gemma-7b-IT output showed hallucinatory content was universally present.

### Evaluation of text similarity and LLM hallucinations

BLEU scores were high for all four task-specific models, reflecting close text similarity post-redaction to the original text (Table 2). This was similar to the BLEU score recorded between clinician redaction and reference text (Table 2). The Levenshtein distances for all task-specific models were low, and slightly lower than the distance reported for clinician redaction, in keeping with high similarity between output and reference texts at the character-level. Qualitative examination of the outputs showed no evidence of hallucination, and therefore that the lower distance likely reflects performance characteristics of the models.

The best performing LLMs, GPT-4-0125 and GPT-3.5-turbo-0125, had consistently similar BLEU scores and Levenshtein distances to values recorded for clinician redaction, and showed no evidence of hallucination on qualitative examination.

Both Phi-3-mini-128k-instruct and Llama-3-8B-Instruct showed improved BLEU scores and Levenshtein distances across zero- and few-shot learning. On qualitative examination, we confirmed that both Phi-3-mini-128k-instruct and Llama-3-8B-Instruct showed evidence of hallucinatory behaviour at zero-, one- and five-shot learning. This included explanations of the task or output (e.g., ‘This text does not contain explicit identifiers, therefore the text remains unchanged’) alongside nonsensical string (e.g., long spans of punctuation). We did not observe any hallucinations at ten-shots.

We report consistently low BLEU scores and high Levenshtein distances across zero- and few-shot learning for Gemma-7b-IT. The output was grossly hallucinatory, including hallucinated medical history (‘Historical factors include prior trauma-related injury sustained one month back’), translations into other languages, and treatment recommendations (‘She’ll have an appointment to see her Dr tomorrow so we can discuss it then’). We therefore did not include a further evaluation of recall for individual PII categories for Gemma-7b-IT.

### PII redaction per category

Names and dates were redacted consistently by all models (Table 3). However, medical record numbers, phone numbers, and the other unique identifiers had variable redaction across models. The Microsoft Azure de-identification service, OBI RoBERTa and BERT models, GPT-4-0125, Llama-3-8B-Instruct and Phi-3-mini-128k-instruct had consistently high recall across PII categories.

**Table 3.**
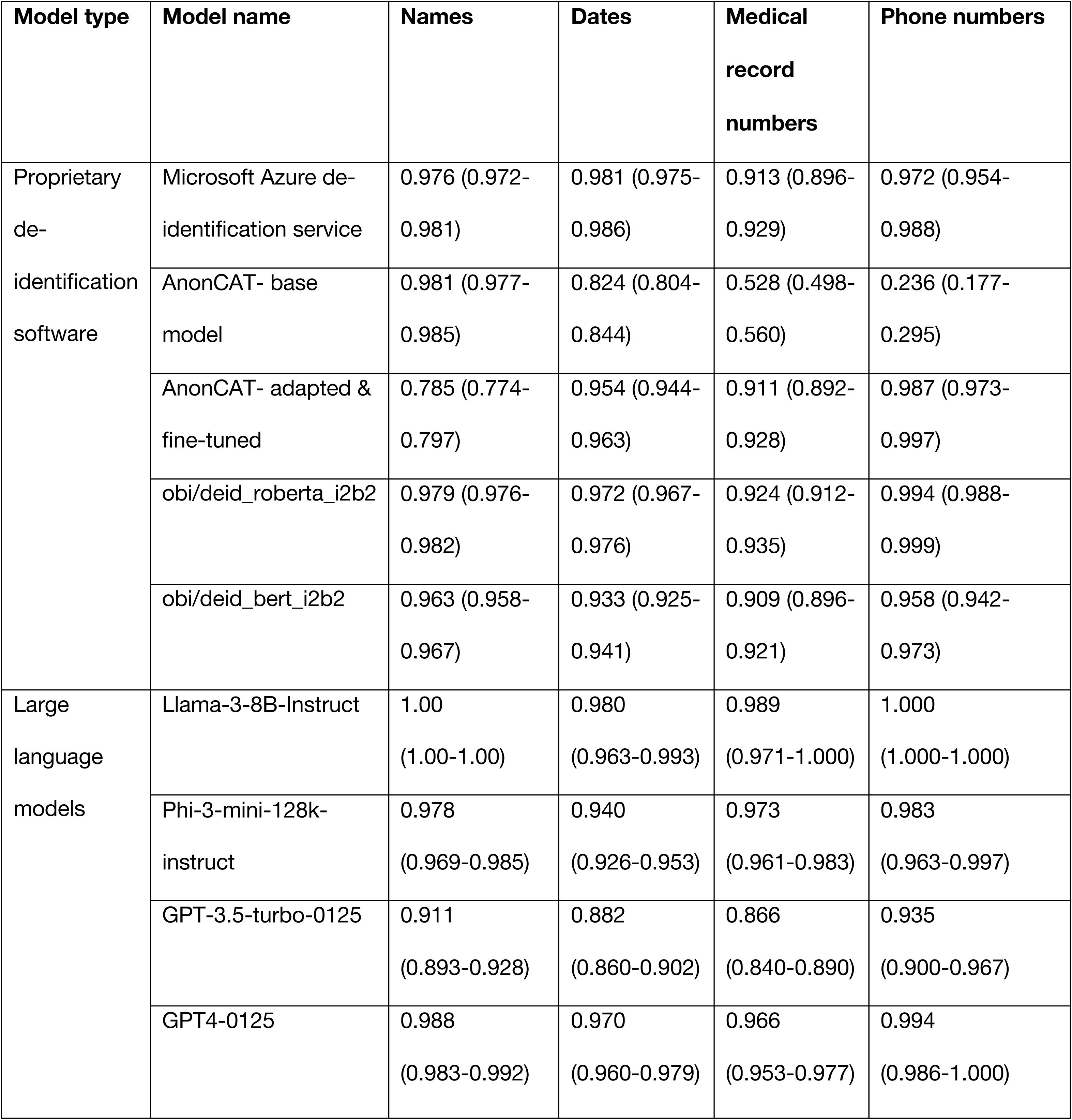
Recall per PII category. Results for AnonCAT are shown for fine-tuning with 365 examples; results for LLMs are shown using 10 shot learning.

| Model type | Model name | Names | Dates | Medical record numbers | Phone numbers |
| --- | --- | --- | --- | --- | --- |
| Proprietary de-identification software | Microsoft Azure de-identification service | 0.976 (0.972-0.981) | 0.981 (0.975-0.986) | 0.913 (0.896-0.929) | 0.972 (0.954-0.988) |
|  | AnonCAT- base model | 0.981 (0.977-0.985) | 0.824 (0.804-0.844) | 0.528 (0.498-0.560) | 0.236 (0.177-0.295) |
|  | AnonCAT- adapted & fine-tuned | 0.785 (0.774-0.797) | 0.954 (0.944-0.963) | 0.911 (0.892-0.928) | 0.987 (0.973-0.997) |
|  | obi/deid_roberta_i2b2 | 0.979 (0.976-0.982) | 0.972 (0.967-0.976) | 0.924 (0.912-0.935) | 0.994 (0.988-0.999) |
|  | obi/deid_bert_i2b2 | 0.963 (0.958-0.967) | 0.933 (0.925-0.941) | 0.909 (0.896-0.921) | 0.958 (0.942-0.973) |
| Large language models | Llama-3-8B-Instruct | 1.00<br>(1.00-1.00) | 0.980<br>(0.963-0.993) | 0.989<br>(0.971-1.000) | 1.000<br>(1.000-1.000) |
|  | Phi-3-mini-128k-instruct | 0.978<br>(0.969-0.985) | 0.940<br>(0.926-0.953) | 0.973<br>(0.961-0.983) | 0.983<br>(0.963-0.997) |
|  | GPT-3.5-turbo-0125 | 0.911<br>(0.893-0.928) | 0.882<br>(0.860-0.902) | 0.866<br>(0.840-0.890) | 0.935<br>(0.900-0.967) |
|  | GPT4-0125 | 0.988<br>(0.983-0.992) | 0.970<br>(0.960-0.979) | 0.966<br>(0.953-0.977) | 0.994<br>(0.986-1.000) |

The base AnonCAT model performed poorly on redacting phone numbers (recall 0.236; 0.177-0.295), but following fine-tuning improved to near-perfect sensitivity (0.987; 0.973-0.997). Qualitative analysis revealed that this was due to differences in the format of internal hospital phone extensions and bleep numbers, which was learned during the tuning process. We observed an unexpected decrease in recall for names after introducing new concepts and performing fine-tuning, possibly indicating some loss of earlier learning (catastrophic forgetting).

### PII redaction per dataset

Of the best performing models, the adapted AnonCAT model showed the least performance shift between dataset, followed by the Microsoft Azure de-identification service (Figure 2, Table S4). GPT-4-0125, the best performing LLM, had a wide range of performance across datasets, with highest performance in the general histopathology dataset, with an F1 0.949 (0.938-0.958), and lowest in the musculoskeletal radiograph dataset, with an F1 0.672 (0.580-0.744). Likewise, GPT-3.5-turbo-0125, Llama-3-8B-Instruct and Phi-3-mini-128k-Instruct had more performance shift across datasets than both task-specific models.

**Figure 2.**
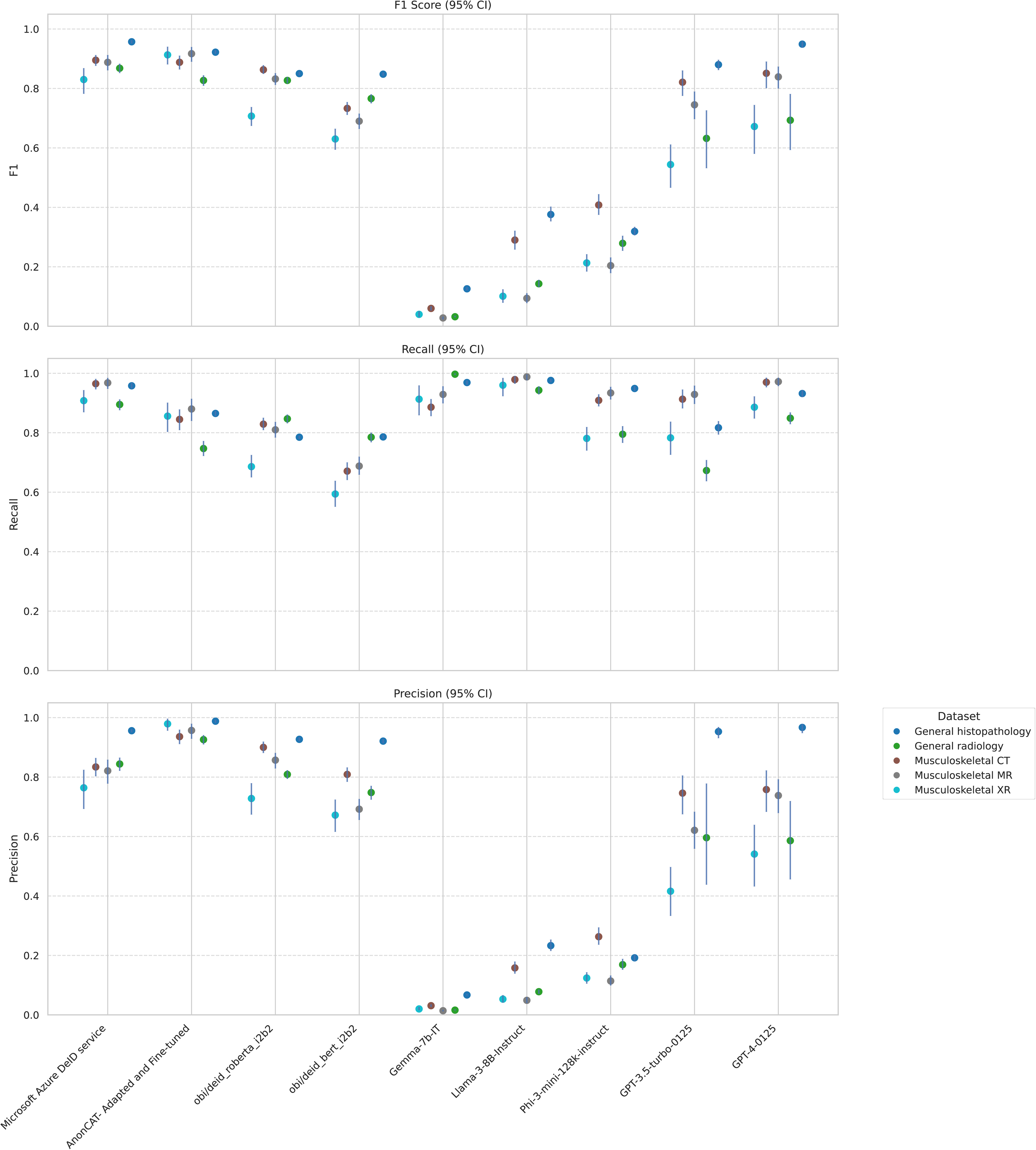
Model performance per dataset, with 95% confidence intervals. All LLM results are plotted using performance with ten-shots. Panel A shows F1 score, panel B recall and panel C precision.

## Discussion

In this study, we evaluated the performance of four task-specific deidentification models and five ‘general-purpose’ LLMs in de-identification of unstructured and semi-structured clinical text, using a dataset of 3650 individual records. Our results suggest that automated de-identification, using either purpose-built software or foundational LLMs, have performance approaching that of human clinicians.

Our findings illustrate that purpose-built redaction models, and LLMs with larger (>8bn) parameter spaces supported by a few in-context examples, provided effective strategies for clinical redaction approaching near-human performance. Where supported, adaptation of models, including fine-tuning of task-specific models and in-context learning of LLMs, commonly improved redaction performance. The Microsoft De-identification service achieved the highest performance in our study, despite not presently supporting site-specific adaptation. The performance of AnonCAT was significantly improved by fine-tuning and addition of concepts to align our study ontology with the model’s default redaction behaviour. This customisable architecture offers significant strengths where a hospital’s redaction needs differ, similar to promptable, generalist LLMs.

The best-performing LLMs in this study de-identified clinical notes with high performance using zero- or few-shot learning, suggesting that it is feasible to avoid or reduce the burden of curating large, manually labelled datasets for model finetuning. However, in all models, we report performance shift across domains. In general, performance was highest in semi-structured histopathology reports, compared to unstructured radiology reports. Task-specific models demonstrated less performance shift across datasets than LLMs.

There was variable performance across LLMs. Both Phi-3-mini-128k-instruct and Llama-3-8B-IT over-redacted records, reflected by consistently high recall in all domains and across all datasets but low precision. Qualitative examination revealed that clinically relevant information was redacted incorrectly; for example, diagnoses or symptoms, reducing the downstream utility of the redacted records. Here, Gemma-7b-IT was particularly prone to hallucination. Qualitative examination showed a mixture of hallucinations, ranging from explaining the task to offering clinical recommendations. This output is not useful for secondary research and may be actively harmful. Increasing LLM parameter size correlated with improved performance, as demonstrated through the superior zero- and few-shot performance characteristics of the GPT-series models.

However, both human and model-based redaction, while good, is not perfect. A single clinician identified 98.6% of all PII, with 96.7% of words redacted representing true PII. The best performing tool overall, Microsoft’s deidentification service identified 95% of PII, including 97.6% of all names, and 92.8% of words redacted represented true PII.

A fundamental issue is what constitutes acceptable performance. Although attractive to strive for perfect redaction, the human performance and inter-annotator variability we describe shows this is unlikely to be possible without significant extra resource. One approach is to examine existing precedent. The MIMIC datasets are amongst the largest publicly available datasets to date. These are redacted by a combination of removing all entries from a database of identifiers, e.g. names, as well as by other string-pattern matching. The recall of this approach on a test corpus was 0.943^10^. Similar performance was achieved by the Microsoft Azure de-identification service, adapted AnonCAT, and GPT-4-0125 in our analysis. Future work could include hybrid approaches based on databases of identifiers, on a population, institution or personal level, to enhance redaction of these alongside generic model-based approaches.

Our study has several limitations. The data from this study derived from a single hospital group in the United Kingdom, and model performance may vary across sites. Our dataset comprises only English-language clinical notes, and we cannot comment on the performance of automated de-identification in other languages. All LLMs in this study were pretrained on an English-dominant corpus, and may transfer differentially to other languages, particularly languages from low-resource settings^31–33^. More extensive adaptation of LLMs such as fine-tuning, as shown for the task-specific AnonCAT, may improve redaction performance particularly where data formats differ significantly between sites. However, this would entail considerable computational expense and technical burden and presently is not amenable to being readily-deployable.

We were unable to evaluate redaction of uncommon, but critical, categories of PII; e.g., email addresses. Furthermore, we were unable to comment on sub-categories of PII, including specific performance for patient names, due to the low prevalence in our dataset. We qualitatively examined 50 examples of output for each model, per-shot, for hallucination, but cannot conclusively report on the prevalence of hallucination for the entire dataset. Finally, we report on seven different comparators, but since conducting the analyses, other software or LLMs may have been developed with superior performance.

If redaction is imperfect, other protections are required to protect privacy and confidentiality. These include requiring researchers to not attempt re-identification of individuals, to only access sufficient data to answer research questions, and retaining data within secure data environments, limiting access to approved researchers. Acceptability of research continues to require multi-stakeholder representation, and is preconditioned on ethical conduct beyond simply de-identification of patient records.

Within the scope of these protections, automating de-identification of clinical notes could enable large-scale sharing of clinical data and its use in training large-scale models, with less time and expense than manual de-identification. Quality assurance through careful examination of model output, across clinical and PII domains, with attention to hallucinatory output and loss of critical information through over-redaction is crucial in real-world practice. Agentic approaches may offer promising avenues for future research, for example, by introducing an ensemble or error-checking mechanisms, with the caveats of increased computational expense.

In summary, our results support the use of automated de-identification systems using no- or low-adaptation strategies. We recommend care and quality assurance in deployment, particularly in implementing systems based on general-purpose LLMs.

## Acknowledgements

This work uses data provided by patients and collected by the UK’s National Health Service as part of their care and support. We thank all the people of Oxfordshire who contribute to the Infections in Oxfordshire Research Database. Research Database Team: L Butcher, H Boseley, C Crichton, DW Crook, DW Eyre, O Freeman, J Gearing (community), R Harrington, K Jeffery, M Landray, A Pal, TEA Peto, TP Quan, J Robinson (community), J Sellors, B Shine, AS Walker, D Waller. Patient and Public Panel: M Ahmed, G Blower, J Hopkins, V Lekkos, R Mandunya, S Markham, B Nichols.

## Data availability

The datasets analysed during the current study are not publicly available as they contain personal data. The histology and general radiology data are available from the Infections in Oxfordshire Research Database (https://oxfordbrc.nihr.ac.uk/research-themes/modernising-medical-microbiology-and-big-infection-diagnostics/iord-about/), subject to an application and research proposal meeting the ethical and governance requirements of the Database. For further details on how to apply for access to the data and a research proposal template please.

## Supplementary methods

### Software and models tested

#### Task-specific deidentification software

##### Microsoft Azure de-identification (DeID) service

The Microsoft Azure DeID service is a paid-for, purpose-built clinical data de-identification pipeline, based on redacting PII using HIPAA categories^1^. The service is accessed within Azure Health Data Services and Microsoft Fabric and can be applied to unstructured or semi-structured text. Evaluation was performed using a private preview version, using the version made available on the 11^th^ April 2024.

##### AnonCAT

AnonCAT is a transformer-based (RoBERTa-large) model, purpose-built for de-identification of unstructured clinical data^2^. It was trained using 2648 manually annotated clinical documents from King’s College Hospital NHS Foundation Trust in the UK. Two smaller sets from Guy’s and St Thomas’ NHS Foundation Trust (328 documents) and University College London Hospitals NHS Foundation Trust (140 documents) were used for fine-tuning and testing the model. AnonCAT is offered as a paid-for service; the core library and annotation tools are open source.

The model version evaluated was 1.1.0 (Model ID: d88707e29606019f).

##### OBI RoBERTa & BERT i2b2 DeID Models

The One Brave IDEA (OBI) RoBERTa and ClinicalBERT DeID models^6^ build upon the respective base transformer models by performing tuning and evaluation using data form the I2B2-DEID dataset and the MassGeneralBrigham (MGB) network. Both models were trained to predict PHI entities within 11 types as defined by HIPAA, and accessed through the Huggingface Transformers API. Evaluation was performed using default parameters.

#### Large language models

##### Gemma-7b-IT

The Gemma models are a family of open-source LLMs, trained on 6T tokens of text, based on Google’s Gemini models^3^. For this study, we evaluated Gemma-7b-IT, which is a 7 billion parameter model.

##### Llama-3-8B-Instruct

The Meta Llama 3 family of LLMs are open-source, auto-regressive language models that use transformer architecture. The model evaluated in this study has 8 billion parameters and is instruction tuned. Information regarding model training and testing are not available via publication.

##### Phi-3-mini-128k-instruct

Phi-3-mini is an open-source, 3.8 billion parameter transformer-decoder model, trained on 3.3 trillion tokens, provided by Microsoft^4^. In this study, we evaluated Phi-3-mini-128k-instruct.

##### GPT-3.5-turbo-0125 and GPT4-turbo-0125

GPT-3.5 and GPT4 are paid-for, transformer based LLMs developed by OpenAI^5^. Detailed information regarding model architecture, training, and testing are not available via publication.

#### LLM hyperparameters

To assess LLM output for zero- and few-shot learning, we used the following hyperparameters: 0, one, five and ten-shots; 3000 was set as the maximum number of output tokens, temperature 0.1, top-k 50 and top-p 0.95. We selected these hyperparameters to increase the predictability and conservativeness of LLM output, given the nature of the task.

OpenAI models (GPT-3.5-turbo-0125 and GPT-4-0125) were used via deployments in the UK South availability zone through the Azure OpenAI service, which is an enterprise grade service that does not retain prompt data for training or service improvement.

#### AnonCAT fine-tuning and adaptation

We fine-tuned AnonCAT using 365 (10%) randomly selected documents from our annotated data, stratified by dataset and presence of patient names within the training sets. We split the fine-tuning dataset into training (292, 80%) and evaluation (73, 20%) sets. Fine-tuning was performed using the Cogstack Modelserve API. As the ontology of AnonCAT de-identification was not based on HIPAA, we added new labels to align the ontology of base-AnonCAT with our labelling ontology. We added concepts for professional titles, external healthcare organisation, hospital/unit, age over 89, account, URL, GMC number, and social security number.

We set a learning rate of 0.00002 over 10 epochs, reporting a final validation loss of 0.05668.

**Figure S1.**
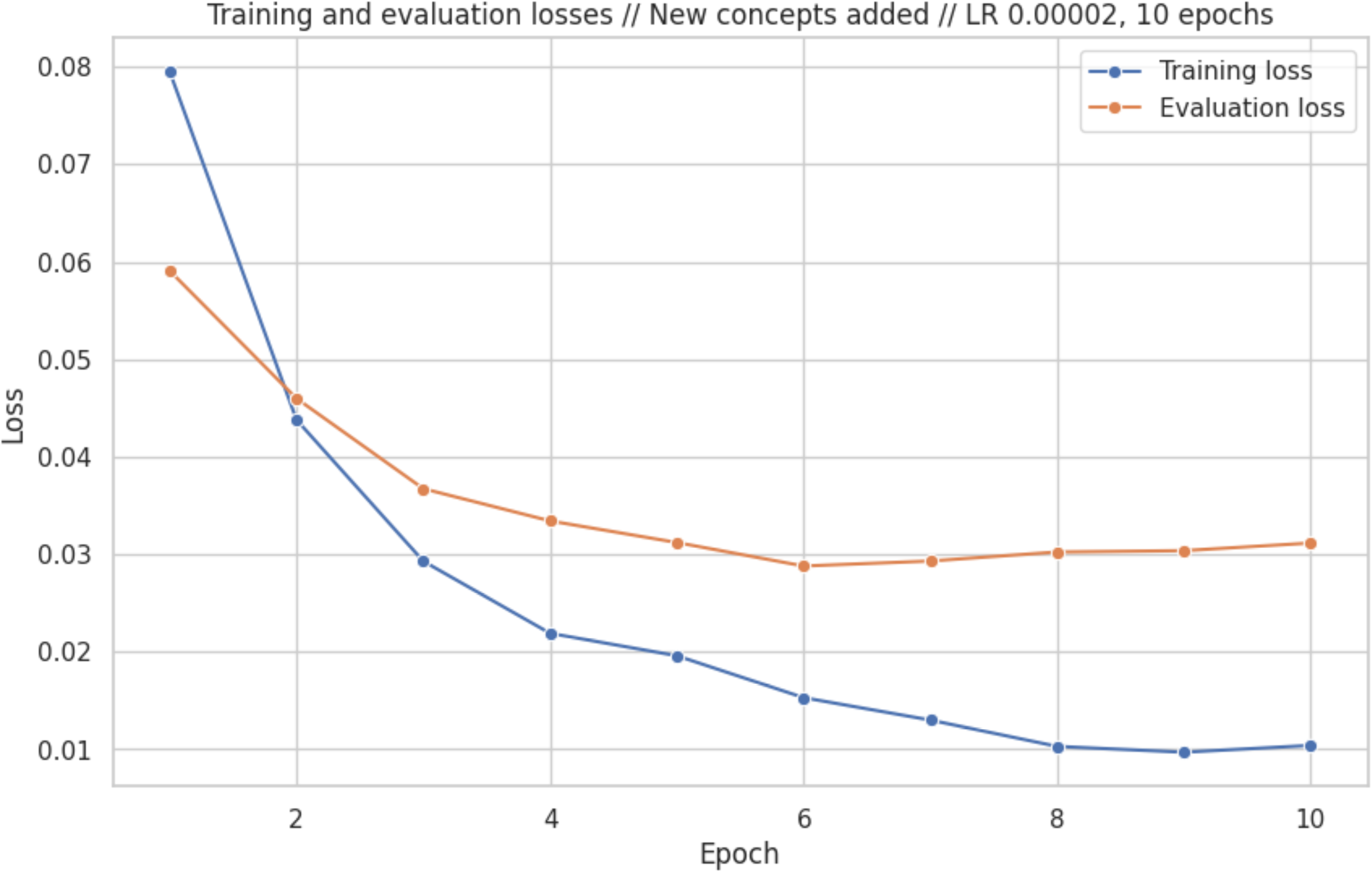
AnonCAT fine-tuning: training and evaluation set loss, over epochs

## Supplementary tables

**Table S1.** Description per dataset

| **Dataset** | **Records** | **Total words** | **Mean words (IQR)** | **Prevalence of PHI (%)** |
| --- | --- | --- | --- | --- |
| Musculoskeletal XR | 550 | 25549 | 46.45 (30-57) | 272 (1.06%) |
| Musculoskeletal CT | 550 | 40920 | 74.40 (44-94) | 688 (1.68%) |
| Musculoskeletal MR | 550 | 55513 | 100.93 (58-132) | 452 (0.81%) |
| General radiology | 1000 | 134385 | 134.49 (61-183) | 1803 (1.34%) |
| General histopathology | 1000 | 223393 | 223.39 (123-262) | 14281 (6.39%) |

**Table S2.** Frequency of PII categories within the dataset

| PII category by frequency | PII subcategory | Count (% of PII) |
| --- | --- | --- |
| Names | **Total** | **6901 (39.44%)** |
|  | Patient or relative | 31 |
|  | Healthcare professional name | 6870 |
| Any other unique identifying number, characteristic, or code | **Total** | **4758 (21.19%)** |
|  | Hospital/unit | 570 |
|  | External healthcare organisation | 1037 |
|  | Professional details | 3151 |
| Dates | **Total** | **3641 (20.81%)** |
|  | Date | 3628 |
|  | Age over 89 years old | 13 |
| Medical record numbers | **Total** | **1408 (8.05%)** |
|  | Medical record number | 22 |
|  | NHS number | 4 |
|  | Specimen identifier | 1382 |
| Phone numbers | **Total** | **334 (1.91%)** |
| Address | **Total** | **133 (0.76%)** |
|  | House or street number | 35 |
|  | Street name | 32 |
|  | City | 27 |
|  | County | 5 |
|  | Country | 18 |
|  | Postcode | 16 |
| Web Universal Resource Locators (URLs) | **Total** | **144 (0.82%)** |
| Certificate/license numbers | **Total** | **107 (0.61%)** |
|  | GMC number | 103 |
|  | NMC number | 0 |
|  | Any other license | 4 |
| Electronic email addresses | **Total** | **59 (0.34%)** |
| Account numbers | **Total** | **10 (0.06%)** |
| Social security numbers | **Total** | **1 (0.01%)** |

**Table S3.** Per model results for classification of PII vs. non-PII.

| Model type | Model name | Number of shots | Precision (95% CI) | Recall (95% CI) | F1 (95% CI) |
| --- | --- | --- | --- | --- | --- |
| **Inter-annotator** |  | **N/A** | **0.967 (0.932-0.993)** | **0.986 (0.971-0.997)** | **0.977 (0.957-0.991)** |
| Task-specific deidentification software | Microsoft Azure de-identification service | N/A | 0.928 (0.922-0.934) | 0.950 (0.943-0.958) | 0.939 (0.934-0.944) |
|  | AnonCAT | No fine-tuning, with requirement to redact professional details | 0.938 (0.932-0.945) | 0.668 (0.660-0.676) | 0.781 (0.774-0.787) |
|  |  | No fine-tuning, without requirement to redact professional details | 0.938 (0.931-0.945) | 0.779 (0.767-0.790) | 0.851 (0.843-0.859) |
|  |  | Adapted and fine-tuned, with requirement to redact professional details | 0.978 (0.973-0.982) | 0.850 (0.843-0.858) | 0.910 (0.905-0.914) |
|  | obi/deid_roberta_i2b2 | N/A | 0.903 (0.898-0.908) | 0.793 (0.789-0.798) | 0.845 (0.841-0.849) |
|  | obi/deid_bert_i2b2 | N/A | 0.880 (0.874-0.887) | 0.775 (0.770-0.780) | 0.824 (0.820-0.829) |
| Large language models | Gemma-7b-IT | 0 | 0.047 (0.045-0.048) | 0.870 (0.859-0.880) | 0.089 (0.086-0.092) |
|  |  | 1 | 0.042 (0.040-0.043) | 0.941 (0.933-0.948) | 0.080 (0.077-0.083) |
|  |  | 5 | 0.043 (0.042-0.045) | 0.974 (0.969-0.978) | 0.083 (0.080-0.086) |
|  |  | 10 | 0.021 (0.019-0.023) | 0.905 (0.885-0.923) | 0.041 (0.037-0.044) |
|  | Llama-3-8B-Instruct | 0 | 0.041 (0.037- 0.045) | 0.943 (0.928- 0.956) | 0.079 (0.072- 0.087) |
|  |  | 1 | 0.048 (0.043-0.052) | 0.984 (0.976-0.991) | 0.091 (0.083-0.010) |
|  |  | 5 | 0.110 (0.099-0.121) | 0.990 (0.983-0.995) | 0.198 (0.181-0.216) |
|  |  | 10 | 0.077 (0.069-0.085) | 0.978 (0.969-0.987) | 0.143 (0.130-0.157) |
|  | Phi-3-mini-128k-instruct | 0 | 0.080 (0.076-0.084) | 0.829 (0.814-0.845) | 0.146 (0.140-0.153) |
|  |  | 1 | 0.155 (0.145-0.166) | 0.820 (0.804-0.835) | 0.261 (0.246-0.276) |
|  |  | 5 | 0.121 (0.115-0.127) | 0.904 (0.892-0.915) | 0.213 (0.204-0.222) |
|  |  | 10 | 0.297 (0.282-0.314) | 0.904 (0.892-0.915) | 0.448 (0.430-0.467) |
|  | GPT-3.5-turbo-0125 | 0 | 0.388 (0.371-0.405) | 0.838 (0.822-0.854) | 0.530 (0.514-0.547) |
|  |  | 1 | 0.476 (0.455-0.496) | 0.767 (0.749-0.784) | 0.588 (0.570-0.604) |
|  |  | 5 | 0.764 (0.725-0.796) | 0.761 (0.745-0.777) | 0.763 (0.741-0.781) |
|  |  | 10 | 0.856 (0.812-0.892) | 0.807 (0.788-0.825) | 0.831 (0.807-0.851) |
|  | GPT-4-0125 | 0 | 0.825 (0.789-0.853) | 0.930 (0.920-0.940) | 0.874 (0.853-0.891) |
|  |  | 1 | 0.814 (0.774-0.848) | 0.931 (0.920-0.941) | 0.868 (0.845-0.888) |
|  |  | 5 | 0.839 (0.797-0.875) | 0.931 (0.921-0.940) | 0.883 (0.858-0.903) |
|  |  | 10 | 0.874 (0.834-0.906) | 0.924 (0.914-0.933) | 0.898 (0.876-0.916) |

**Table S4.**
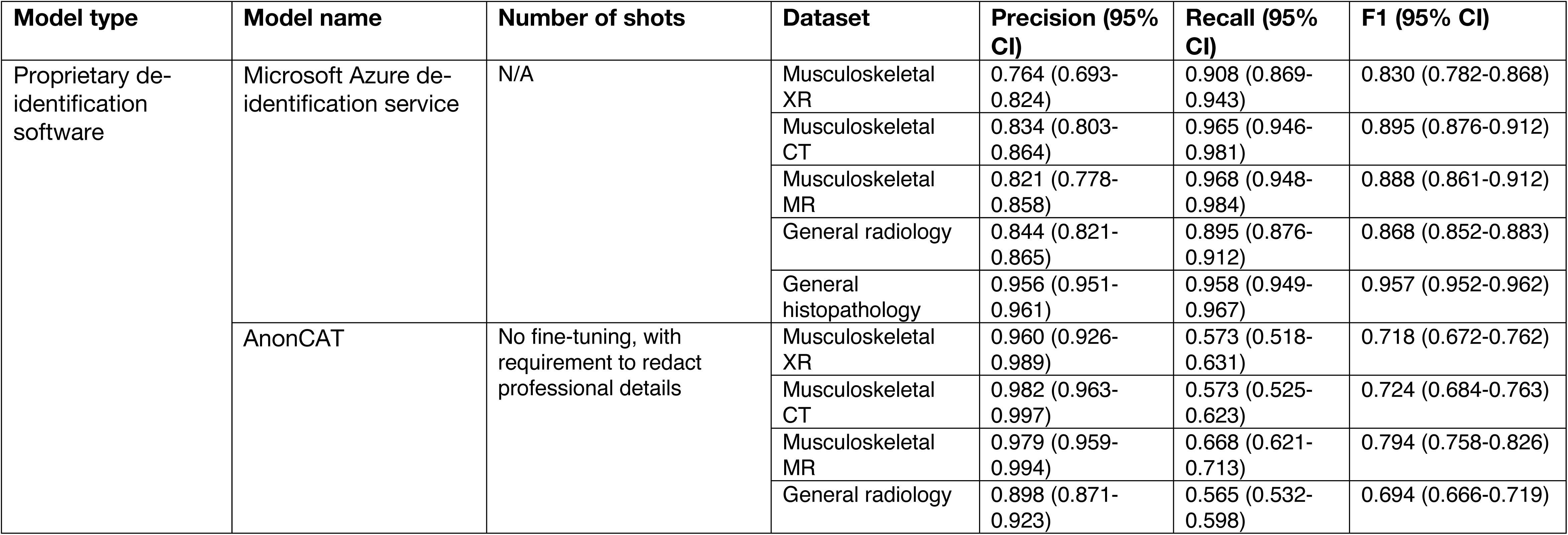
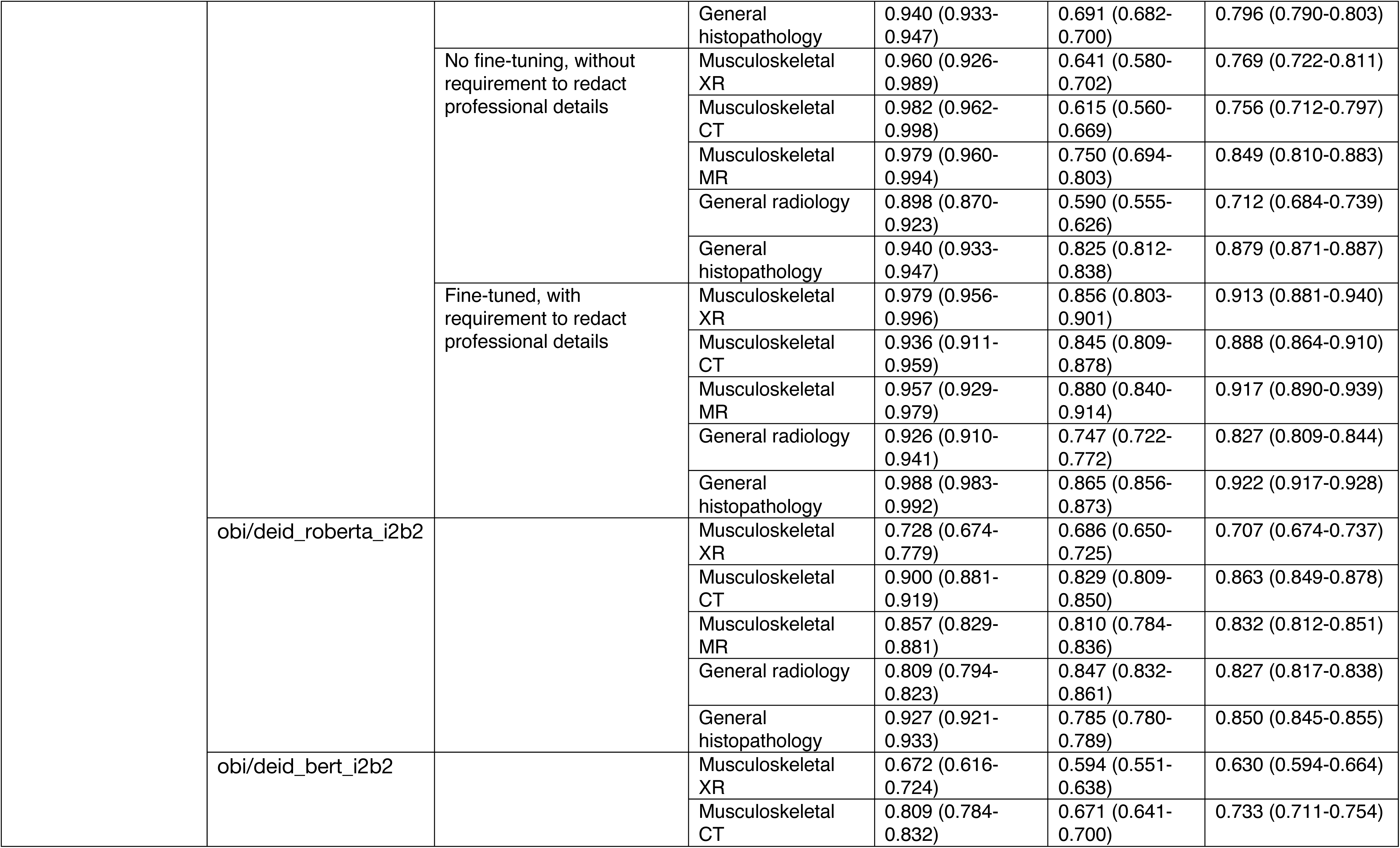
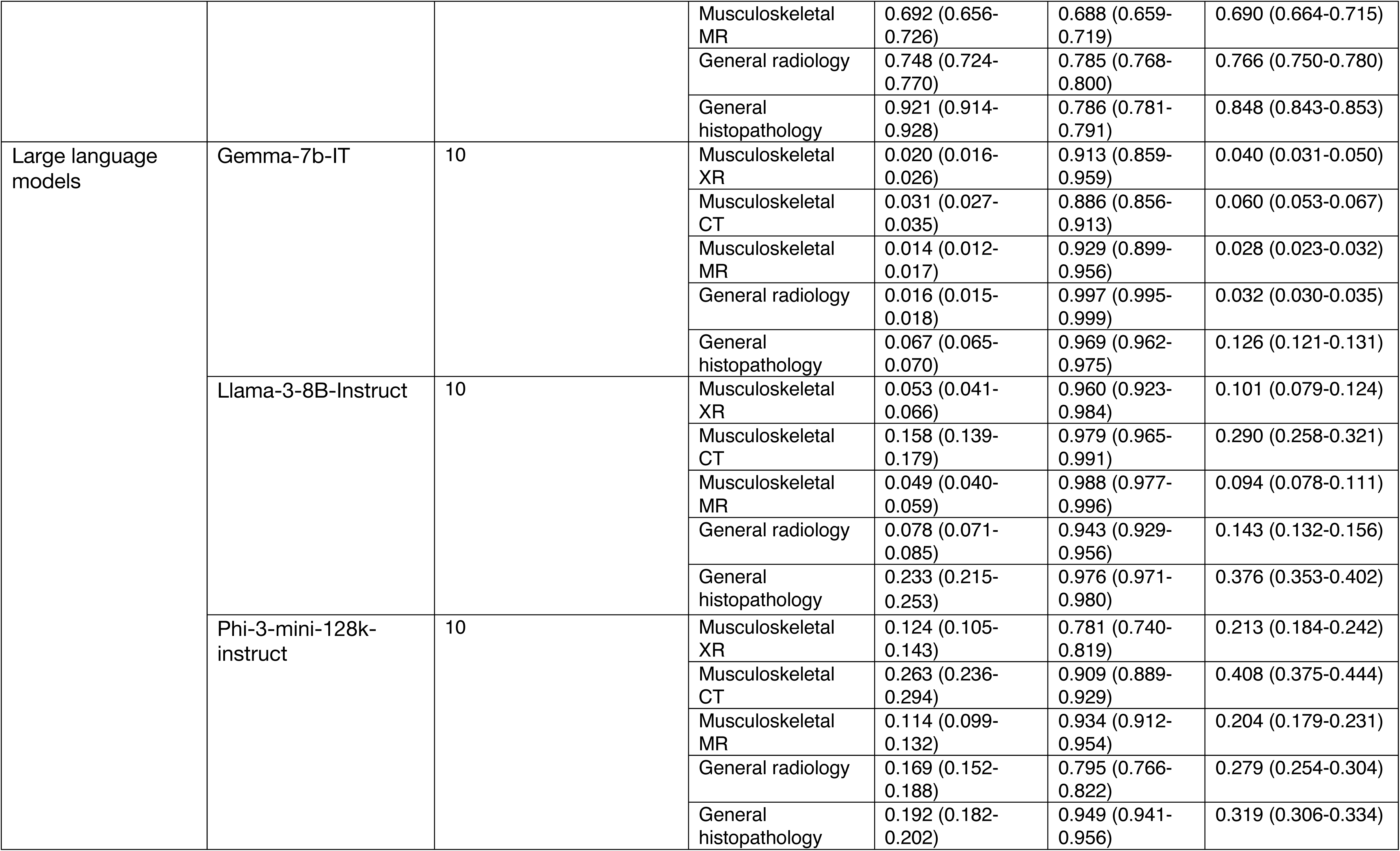
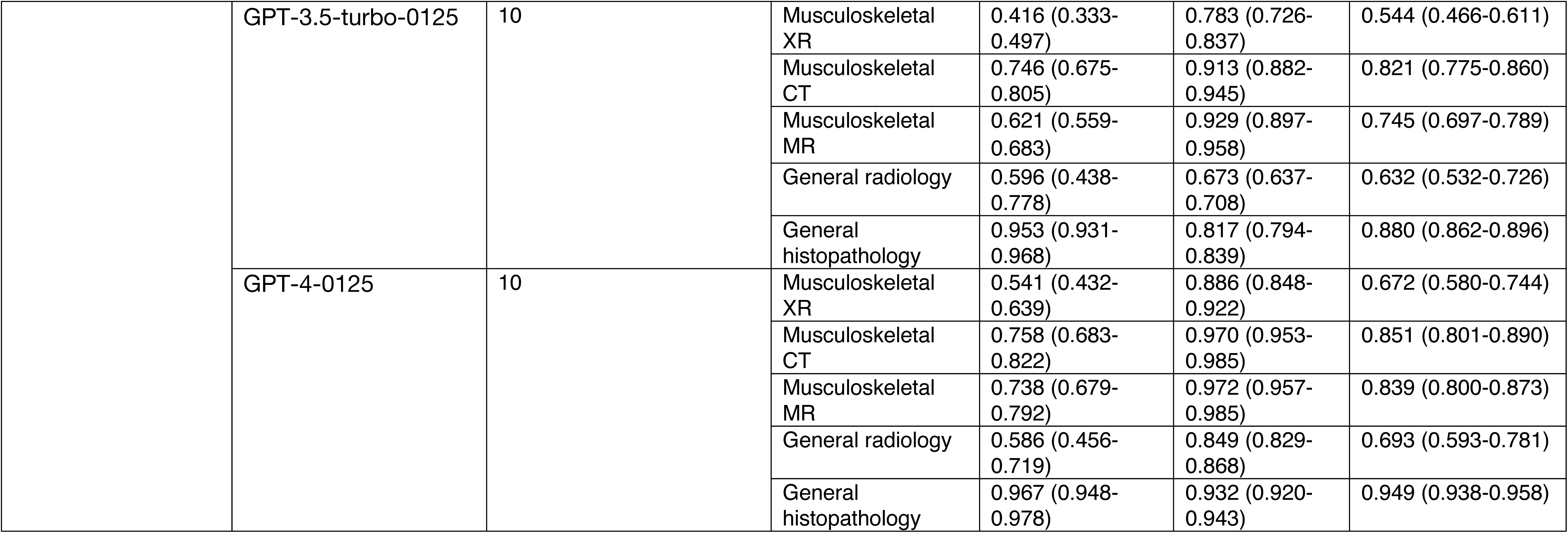
Model precision, recall and F1 score per dataset

## Supplementary figures

**Figure S1.**
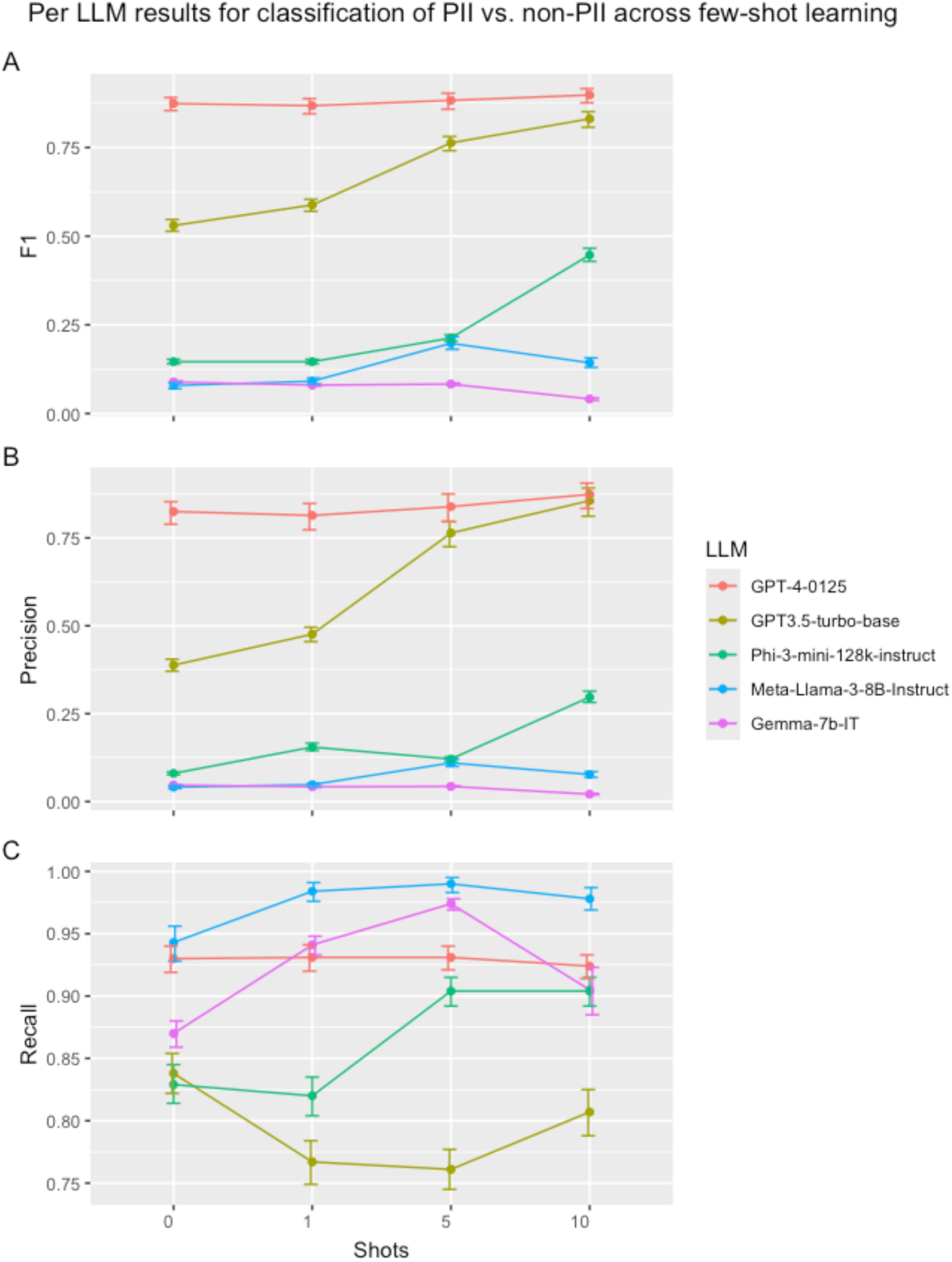
LLM performance by shot

## References

1. Henry J, Pylypchuk Y, Searcy T, Patel V. Adoption of electronic health record systems among US non-federal acute care hospitals: 2008–2015. ONC data brief 2016;35(35):2008–2015.

2. Slawomirski L, Lindner L, de Bienassis K, Haywood P, Hashiguchi TCO, Steentjes M, et al. Progress on implementing and using electronic health record systems: Developments in OECD countries as of 2021. 2023.

3. Shah SM, Khan RA. Secondary use of electronic health record: Opportunities and challenges. 2020;8:136947–136965.

4. Xiao C, Choi E, Sun J. Opportunities and challenges in developing deep learning models using electronic health records data: a systematic review. 2018;25(10):1419–1428.

5. Kalkman S, van Delden J, Banerjee A, Tyl B, Mostert M, van Thiel G. Patients’ and public views and attitudes towards the sharing of health data for research: a narrative review of the empirical evidence. J.Med.Ethics 2022;48(1):3–13.

6. Kuo RYL, Freethy A, Smith J, Hill R, Joanna C, Jerome D, et al. Stakeholder perspectives towards diagnostic artificial intelligence: a co-produced qualitative evidence synthesis. 2024;71.

7. Computer-assisted de-identification of free text in the MIMIC II database. Computers in Cardiology, 2004: IEEE; 2004.

8. Steinkamp JM, Pomeranz T, Adleberg J, Kahn Jr CE, Cook TS. Evaluation of automated public de-identification tools on a corpus of radiology reports. 2020;2(6):e190137.

9. Kovačević A, Bašaragin B, Milošević N, Nenadić G. De-identification of clinical free text using natural language processing: A systematic review of current approaches. Artif.Intell.Med. 2024:102845.

10. Neamatullah I, Douglass MM, Lehman LH, Reisner A, Villarroel M, Long WJ, et al. Automated de-identification of free-text medical records. 2008;8:1–17.

11. Instruction-guided deidentification with synthetic test cases for Norwegian clinical text. Northern Lights Deep Learning Conference: PMLR; 2024.

12. Vaswani A, Shazeer N, Parmar N, Uszkoreit J, Jones L, Gomez AN, et al. Attention is all you need. 2017;30.

13. Labrak Y, Rouvier M, Dufour R. A zero-shot and few-shot study of instruction-finetuned large language models applied to clinical and biomedical tasks. 2023.

14. Liu Z, Huang Y, Yu X, Zhang L, Wu Z, Cao C, et al. Deid-gpt: Zero-shot medical text de-identification by gpt-4. 2023.

15. Xu Z, Jain S, Kankanhalli M. Hallucination is inevitable: An innate limitation of large language models. 2024.

16. Huang L, Yu W, Ma W, Zhong W, Feng Z, Wang H, et al. A survey on hallucination in large language models: Principles, taxonomy, challenges, and open questions. 2023.

17. Rawte V, Sheth A, Das A. A survey of hallucination in large foundation models. 2023.

18. OUHNFT. Oxford University Hospitals NHS Foundation Trust. Available at: https://www.ouh.nhs.uk/about/. Accessed June, 2024.

19. Act A. Health insurance portability and accountability act of 1996. 1996;104:191.

20. Hironsan. doccano: an open-source text annotation tool. Accessed June, 2024.

21. Mavon K. Microsoft De-Identification Service.. Accessed June, 2024.

22. Validating transformers for redaction of text from electronic health records in real-world healthcare. 2023 IEEE 11th International Conference on Healthcare Informatics (ICHI): IEEE; 2023.

23. Team G, Mesnard T, Hardin C, Dadashi R, Bhupatiraju S, Pathak S, et al. Gemma: Open models based on gemini research and technology. 2024.

24. Abdin M, Jacobs SA, Awan AA, Aneja J, Awadallah A, Awadalla H, et al. Phi-3 technical report: A highly capable language model locally on your phone. 2024.

25. Liu Q, Hyland S, Bannur S, Bouzid K, Castro DC, Wetscherek MT, et al. Exploring the Boundaries of GPT-4 in Radiology. 2023.

26. Wang J, Shi E, Yu S, Wu Z, Ma C, Dai H, et al. Prompt engineering for healthcare: Methodologies and applications. 2023.

27. Hripcsak G, Rothschild AS. Agreement, the f-measure, and reliability in information retrieval. 2005;12(3):296–298.

28. Yujian L, Bo L. A normalized Levenshtein distance metric. IEEE Trans. Pattern Anal.Mach.Intell. 2007;29(6):1091–1095.

29. Bleu: a method for automatic evaluation of machine translation. Proceedings of the 40th annual meeting of the Association for Computational Linguistics; 2002.

30. Steinkamp JM, Pomeranz T, Adleberg J, Kahn Jr CE, Cook TS. Evaluation of automated public de-identification tools on a corpus of radiology reports. 2020;2(6):e190137.

31. Zhao J, Zhang Z, Zhang Q, Gui T, Huang X. Llama beyond english: An empirical study on language capability transfer. 2024.

32. Li Z, Shi Y, Liu Z, Yang F, Liu N, Du M. Quantifying Multilingual Performance of Large Language Models Across Languages. 2024.

33. Achiam J, Adler S, Agarwal S, Ahmad L, Akkaya I, Aleman FL, et al. Gpt-4 technical report. 2023.

## Supplementary references

1. Mavon K. Microsoft De-Identification Service. https://techcommunity.microsoft.com/t5/healthcare-and-life-sciences/announcing-a-de-identification-service-for-health-and-life/ba-p/3949712 Web site. Accessed June, 2024

2. Kraljevic Z, Shek A, Yeung JA, et al. Validating transformers for redaction of text from electronic health records in real-world healthcare. 2023 IEEE 11th International Conference on Healthcare Informatics (ICHI)

3. Team G, Mesnard T, Hardin C, et al. Gemma: Open models based on gemini research and technology. *arXiv preprint arXiv:2403.08295.* 2024

4. Abdin M, Jacobs SA, Awan AA, et al. Phi-3 technical report: A highly capable language model locally on your phone. *arXiv preprint arXiv:2404.14219*. 2024

5. Achiam J, Adler S, Agarwal S, et al. Gpt-4 technical report. *arXiv preprint arXiv:2303.08774.* 2023

6. Kailas P, Goto S, Homilius M, MacRae CA, Deo RC. obi-ml-public/ehr_deidentification. Zenodo. 2022.

